# The Effect of Single Pulse TMS on Working Memory Performance: An Online TMS And fNIRS Study

**DOI:** 10.64898/2026.07.22.26358678

**Authors:** Summer Edwards, Quinn Smith, Jesse Farrand, Tressie M. Stephens, Lei Ding, Andrew K. Conner, Ian F. Dunn, Mark S. George, Han Yuan

**Affiliations:** Stephenson School of Biomedical Engineering, The University of Oklahoma, Norman, OK, USA; Department of Neurosurgery, University of Oklahoma Health Campus, Oklahoma City, OK, USA; Institute for Biomedical Engineering, Science and Technology, The University of Oklahoma, Norman, OK, USA; Department of Psychiatry and Behavioral Sciences, Medical University of South Carolina, Charleston, SC, USA; Ralph H. Johnson VA Medical Center, Charleston, SC, USA

**Keywords:** Transcranial magnetic stimulation, functional near-infrared spectroscopy, working memory, dorsolateral prefrontal cortex, online TMS paradigm

## Abstract

Transcranial magnetic stimulation (TMS) is widely used in both clinical and research settings to study and treat neurological and neuropsychiatric disorders, yet its underlying neural mechanisms remain unclear; particularly how stimulation influences both local and distant regional activity in relation to behavior. In this study, we combined single-pulse TMS with concurrent whole-head functional near-infrared spectroscopy (fNIRS) to examine hemodynamic and behavioral responses during a working memory task, with a focus on behavioral variability. Single TMS pulses were delivered to the left dorsolateral prefrontal cortex (DLPFC) while healthy participants rested; additionally, single pulses were delivered online to the left DLPFC while participants performed the working memory task. Across all participants, we observed a reliable load-dependent increase in hemodynamic activity associated with task. However, behavioral responses to TMS varied during the task. When participants were stratified into subgroups based on performance, a distinct topographic pattern emerged. During the task, TMS systematically modulated hemodynamic responses in regions including DLPFC, superior medial gyrus, precuneus, and parietal lobule, which are areas belonging to the default mode network. Moreover, the hemodynamic response during the single pulse alone sessions without any task was also found to be associated with the behavioral responses in a coherent pattern involving DLPFC, precuneus and parietal lobules. These findings suggest that variable behavioral outcomes during online TMS task are linked to distinct hemodynamic responses in a topographic pattern of local and distant regional areas.

## Introduction

Transcranial magnetic stimulation (TMS) is a method used to excite or inhibit neurons through single or repetitive pulses (Lozano and Hallett, 2013). In the past decade, there have been many advances to TMS use as FDA-approved treatment for major depressive disorder, obsessive compulsive disorder, migraine associated pain, and smoking cessation (Cohen et al., 2022; Perera et al., 2016). It is also clinically used for brain tumor mapping, and stroke rehabilitation (Fisicaro et al., 2019; Krieg, 2017). However, these approaches have been variably successful (Carmi et al., 2018; Carpenter et al., 2012; Lefaucheur et al., 2020; Lipton et al., 2010; Perera et al., 2016). In depression treatment for example, only 15-30% of patients reach remission in double-blind trials, or 30-50% remission in open label studies using TMS (Sonmez et al., 2019). Thus, there is a need to better understand what drives the response variability.

Although TMS can modulate brain activity, response variability hinders its clinical and research usage. A key strategy for addressing this challenge is to measure target engagement, particularly with concurrent functional imaging (Sack et al., 2024). Target engagement is the extent to which intended brain regions are being effectively modulated through measurable changes, and is evidence of spatial accuracy, physiological effect, and functional relevance. Target engagement demonstrates that TMS stimulation actually reaches and modulates the “targeted” brain region or network and produces the predicted neurophysiological or behavioral effect (Sack et al., 2024). The most direct physiological marker includes movement or motor evoked potential from motor cortex stimulation, like thumb twitching from resting state motor threshold; however, cognitive and network-level measures can be acquired through concurrent assessment and imaging. In the treatment of depression, target engagement can be verified behaviorally and biologically: behaviorally through symptom reduction, and biologically through measured hemodynamic, functional, evoked potential, or plasticity changes (Dubin, 2017; Philip et al., 2018a).

Recently, an intriguing way to probe target engagement is through cognitive tasks that directly tap the neural circuits implicated in depression. Working memory has emerged as a sensitive measure, as engaging these processes can both reflect and potentially enhance the fronto-parietal network as the target of treatment (Bradley et al., 2022; Feredoes et al., 2011; Sack et al., 2024; Sathappan et al., 2019; Zokaei et al., 2014). Working memory tasks, such as the N-back task, consistently engage regions like the dorsolateral prefrontal cortex (DLPFC) in a load-dependent manner, and have easily interpretable behavior results (Owen et al., 2005). This provides a reproducible baseline against which TMS-induced changes could be measured; especially since TMS pulses are approved for DLPFC delivery.

The behavioral relationship between TMS and working memory function has been investigated in many studies with various designs or conflicting reports (Brunoni and Vanderhasselt, 2014; Haque et al., 2021; Tozzi et al., 2024). In healthy adults, single pulse TMS applied over the left DLPFC during a 3-back task resulted in increased errors compared to no TMS controls; TMS applied over the right DLPFC did not alter working memory performance (Mull and Seyal, 2001). Additionally, during a verbal working memory task, double-pulse TMS to the left DLPFC between conditions also decreased performance in healthy adults (Osaka et al., 2007). Repetitive TMS at 10 Hz to the right DLPFC can have the opposite effect in clinical groups during the N-back task, however (Hulst et al., 2017). One specific study shows that this design does in fact improve N-back performance in persons with multiple sclerosis compared to sham TMS (Hulst et al., 2017), but has no significant effects in healthy controls (Hulst et al., 2017; Grosshagauer et al., 2025). These mixed behavioral effects are like those mentioned in clinical effectiveness: TMS can enhance or disrupt working memory in healthy persons and pose therapeutic or no-change in clinical populations.

Hulst et al. additionally collected offline functional imaging data and found that task-related frontal activation following repetitive TMS correlated with clinical group N-back performance (2017). This supports the concept of assessing behavioral variability by combining a working memory task with TMS and functional imaging. Other studies, however, have used *online* designs using interleaved functional magnetic resonance imaging (fMRI) and TMS to probe working memory. For example, Webler et al. applied single-pulse TMS to the left DLPFC during an N-back task while acquiring concurrent fMRI data. Their results showed that single-pulse stimulation to the left DLPFC improved working memory accuracy in healthy participants and activated the intended target regions (2022). However, this approach has several drawbacks, including high cost and challenges for clinical usage, as fMRI cannot be used with subjects who have metal implants or experience claustrophobia. Additionally, most clinics lack the resources for concurrent fMRI-TMS, as the setup requires a highly controlled environment.

Functional near infrared spectroscopy (fNIRS) could be an alternative to fMRI-TMS. fNIRS emits near-infrared light through the scalp and is absorbed by hemoglobin in the blood. These absorptions can be quantified into concentration changes of oxygenated and deoxygenated hemoglobin (HbO and HbR) through established preprocessing pipelines (Zhang et al., 2021). In combination with TMS, there is no magnetic interference with pulse delivery, allowing for fully concurrent recording, efficient setup, more feasibility and cost-saving with fNIRS.

Our recent work showed feasibility of simultaneous fNIRS-TMS with electroencephalography (EEG), namely fNET (Farrand et al., 2025). The current study used whole-head, simultaneous fNIRS and TMS imaging to investigate the online effects of TMS on working memory *for the first time*. We acquired whole-head fNIRS while subjects performed a 0-Back and 2-Back (namely low-load and high-load) working memory task with and without TMS, and examined the working memory, imaging, and TMS relationships. It was hypothesized that the fNIRS recorded hemodynamic responses would measure those brain areas engaged by the working memory task and TMS, and that TMS would perturb working memory behavior and imaging measures. In addition, we also acquired fNIRS recordings when subjects received single pulse TMS pulses alone without any task. We examined whether the hemodynamic response to TMS without the working memory task are intrinsically related to the behavioral responses during the task.

## Materials and Methods

### Participants

All study protocols were approved by the Institutional Review Board at The University of Oklahoma Health Sciences Center (OUHSC) under IRB No. 9915. The research was conducted in accordance with the principles embodied in the Declaration of Helsinki and in accordance with local statutory requirements. All participants gave written informed consent to participate in the study. Right-handed healthy adults were recruited for this study and signed written consent before any experimental procedures. Subjects were screened for neurological or neuropsychiatric diseases, MRI safety, TMS safety, sleep disorder, and pregnancy; if any were present, they were excluded from the study. Additionally, all subjects were fluent in English, completed at least 10 years of education, had normal hearing and vision, and were non-smoking.

### Experiment and Task Design

The following tasks were completed in every subject: resting motor threshold (RMT), resting state 1, working memory 1, working memory 2, working memory 3, single pulse, motor, resting state 2. All tasks were completed at 2 recording visits spaced about one month apart; an additional MRI visit was completed prior to any recording visits.

### Data Acquisition

Simultaneous fNIRS, EEG, and peripheral measurements of pulse oximetry, accelerometry, and respiration were recorded in every subject with the use of a bite bar to control jaw movements, following prior recording setups (Chen et al., 2020; Zhang et al., 2021; Zhang et al., 2023). 32 source probes, 31 detector probes and 8 short-separation detectors were arranged over the whole head totaling 110 channels (i.e. 110 pairs of sources and detectors). Among these, 8 short-separation channels were evenly spread throughout the head. Signals were acquired at a sampling rate of 7.81 Hz. To ensure good quality, fNIRS were calibrated to have good or excellent quality in ≥ 90% of all channels. EEG measurements were acquired by a Brain Vision actiCHamp system (BrainProducts, München, Germany). 64 EEG channels were spaced evenly throughout the head following the international 10-5 system. The electrode at Pz position was selected as the reference while ground was placed at AFp1. Electrically conductive gel was used with impedance of EEG electrode under 20 kW. EEG data was acquired at a sampling rate of 5000 Hz but was *not* included in the current report.

TMS was delivered using Magventure MagPro X100 and Cool-B65 coil (MagVenture, Inc, Georgia, USA) system. Custom E-Prime 2.0 (Psychology Software Tools, PA, USA) controlled the timing TMS pulse delivery and task trials; only working memory and single pulse session included TMS pulses. Motor threshold was determined using PEST (Borckardt et al., 2006), while monitoring right-hand thumb twitches during motor cortex stimulation with the coil oriented at 45°. TMS pulses were delivered to the left DLPFC at 100% resting motor threshold, which was targeted using BEAM-F3 method (Beam et al., 2009). The Cool-B65 coil was placed parallel to the defined target with a Magventure Super Flex Arm (MagVenture, Inc, Georgia, USA). The online TMS working memory paradigm is illustrated in *Figure 1.* Each participant performed the online TMS working memory task three times per visit. During working memory, there were a total of 12 N-back blocks consisting of 12 letter trials per run, repeated 3 times. The order of blocks was as follows: low-load (0-back), low-load with TMS (0-back-TMS), high-load (2-back), high-load with TMS (2-back-TMS), low-load with TMS, low-load, high-load with TMS, high-load, low-load, high-load, low-load with TMS, high-load with TMS. The blocks with TMS had a total of 7 pulses distributed during letter presentation with an inter-pulse interval of 2.4 s, totaling 126 pulses per recording. The timing of letter presentation and TMS pulses followed the protocol used in Webler et al., which was designed to align with MRI repetition time constraints (2022). In addition to the online TMS working memory task, each subject had a single pulse alone session once per visit. During the single pulse alone session, 46 pulses were delivered and the inter pulse interval was pseudorandomized and uniformly distributed from 6.4 to 9.6 seconds, following the timing protocol used in Pantazatos et al. (2023).

**Fig. 1.**
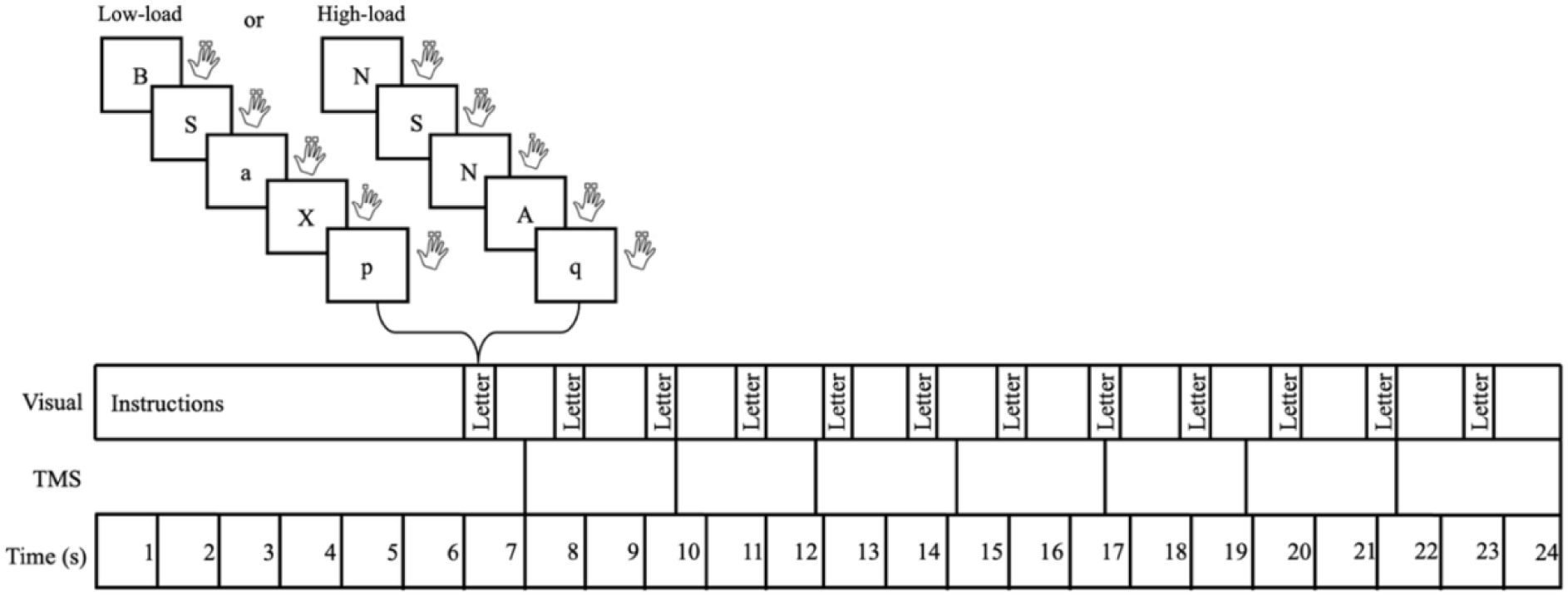
Diagram of online-TMS working memory paradigm. All letters presented would either be low-load or high-load, with or without TMS. This figure illustrates the timing of TMS pulses during blocks with TMS.

Subjects had MRI scans on a GE Signa 3T Architect scanner at OUHSC. While structural, functional, and diffusion MRIs were collected, only the T1 MRI was used for this study to complete co-registration between MRI and fNIRS sensors. Localite TMS Navigator (Localite GmBH, Germany) frameless stereotaxy system was used for digitizing the sensor positions.

### Behavior Data Analysis

Responses and reaction time of working memory tasks were output on E-prime files. The calculation of performance accuracy and statistical analysis were performed using Excel and MATLAB^®^. Average accuracy during low-load and high-load conditions were calculated for each condition: no TMS and TMS. Accuracy measures include both incorrect and missed responses, while reaction time only includes correct responses. There were no behavior outputs from the single-pulse TMS session.

### fNIRS Data Analysis

fNIRS data analysis was completed in the following steps: quality assessment, preprocessing, and statistical parameter mapping. Quality assessment was completed on nirsLAB_v201 to visually inspect data prior to motion correction and short channel regression. Then, data was preprocessed by adapting an automatic denoising procedure, principal-component-analysis-based general linear model (PCA-GLM) (Zhang et al., 2021). The preprocessing steps include rejecting bad channels that showed no heartbeat frequency peak, identifying and rejecting bad time segments with excessing head movements, band-pass filtering, and removing the physiological noises of superficial contribution, respiration, cardiac pulsation, and motion acceleration recorded by EEG and peripheral measurements. Cleaned data are then processed in NIRS_SPM_v4, so that statistical analyses at the sensor level can be projected onto a rendered brain surface for visualization (Li et al., 2012; Ye et al., 2009). NIRS_SPM is set with no pre-whitening, a basis function of hemodynamic response function, discrete cosine transform temporal filtering, no pre-coloring, and no serial correlation corrections. Each subject is processed individually with their digitization coordinates that were co-registered with their T1, transformed to MNI (Montreal Neurological Institute) coordinates, and projected onto the standard rendered brain space. Group analysis used Lipschitz-Killing curvature based expected Euler characteristics for the multiple comparison correction of *p* values (Li et al., 2012).

For the working memory task data, the events of low-load and high-load with and without TMS were modeled using the general linear model (GLM) analysis. The load effect was estimated as contrasting the high-load and low-load conditions across TMS and no TMS conditions, i.e., 2-back-TMS + 2-back– 0-back-TMS – 0-back. The TMS effect was estimated as contrasting working memory task with TMS and working memory task without TMS conditions, i.e., 2-back-TMS + 0-back-TMS – 2-back – 0-back. In the stratified analysis, subjects were separated into subgroups of improved, decreased and no change subgroups based on their behavioral performance during TMS. The improved category was defined when TMS increased the accuracy of high-load condition, while the decreased and no change categories had respective responses to TMS. ANOVA was performed based on behavioral stratification to identify the areas associated with the TMS effect. In this ANOVA analysis, the independent variable was the subgroup category, and the dependent variable was the estimated HbO change of the TMS effect (i.e., 2-back-TMS + 0-back-TMS – 2-back – 0-back). Regions of interest (ROI) was defined on the group map of the ANOVA analysis, then post-hoc analysis was performed using pairwise *t* test and Bonferroni correction was used for multiple comparison correction.

Furthermore, to explore any potentially intrinsic association of hemodynamic response to TMS pulses alone and the behavioral responses from a separate task, ANOVA analysis of behavioral stratification (increased, decreased and no change) was performed on fNIRS recordings during the single pulse session without the working memory task. This ANOVA analysis was set the same where the independent variable was the subgroup category, and the dependent variable was the estimated hemodynamic response of the single pulse TMS effect (i.e., single pulse on vs. the rest). Regions of interest (ROI) was defined on the group map of the ANOVA analysis, then post-hoc analysis was performed using pairwise *t* test and Bonferroni correction was used for multiple comparison correction.

## Results

### Demographic and Behavior Data

22 healthy subjects were recruited in total; however, 2 were immediately withdrawn due to scheduling conflict or cap intolerability. Thus, 20 subjects participated in the study and their demographics are listed in *Table 1.* The subjects aged from 21 to 49 years, and all subjects completed high school and college education. Slightly fewer subjects (N = 17) had complete data from both the working memory and single pulse sessions and are included in the current report.

**Table 1:** Demographics.

| Subject<br>(N = 20) | Age<br>(Years) | Gender<br>(Female/Male/<br>Unknown) | Ethnicity<br>(Hispanic/Non-Hispanic/Unknown) | Education<br>(Years) |
| --- | --- | --- | --- | --- |
| Mean | 33.25 | 13/7/0 | 1/18/1 | 16.85 |
| STE | 2.06 | - | - | 0.59 |

Behavioral results are listed in *Table 2.* Whole-group performance appeared to have non-significant change by TMS, in terms of both accuracy and reaction time (illustrated in *Figure 2*). Variable performances are observed, so the data were split into improved, decreased, and no change subgroups. The stratified accuracies across subgroups are shown in *Figure 3,* where changes between improved and decreased subgroups are significant (*p* < 0.001). However, neither the years of education nor the age of subjects was associated with the stratified performance (*p* >= 0.1).

**Fig. 2.**
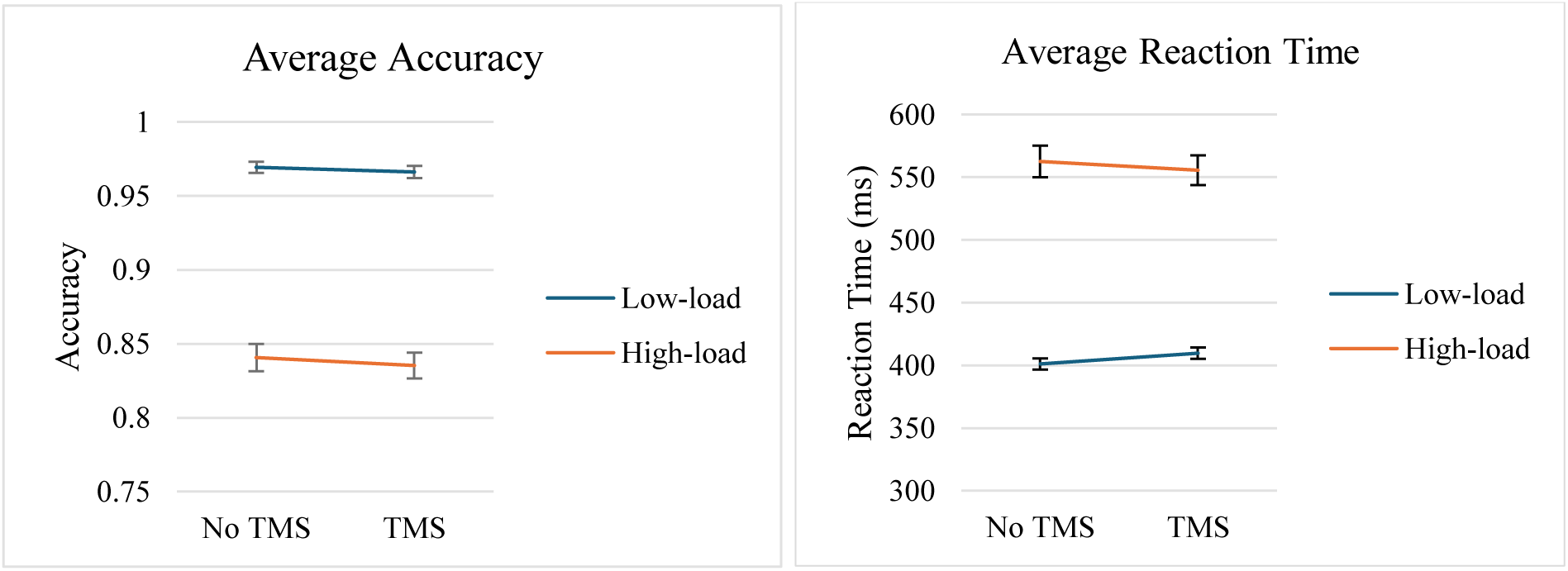
Average accuracy and reaction time during low-load and high-load conditions across all subjects. There is no significant difference in accuracy or reaction time by TMS at whole-group level. Bars represent standard error.

**Fig. 3.**
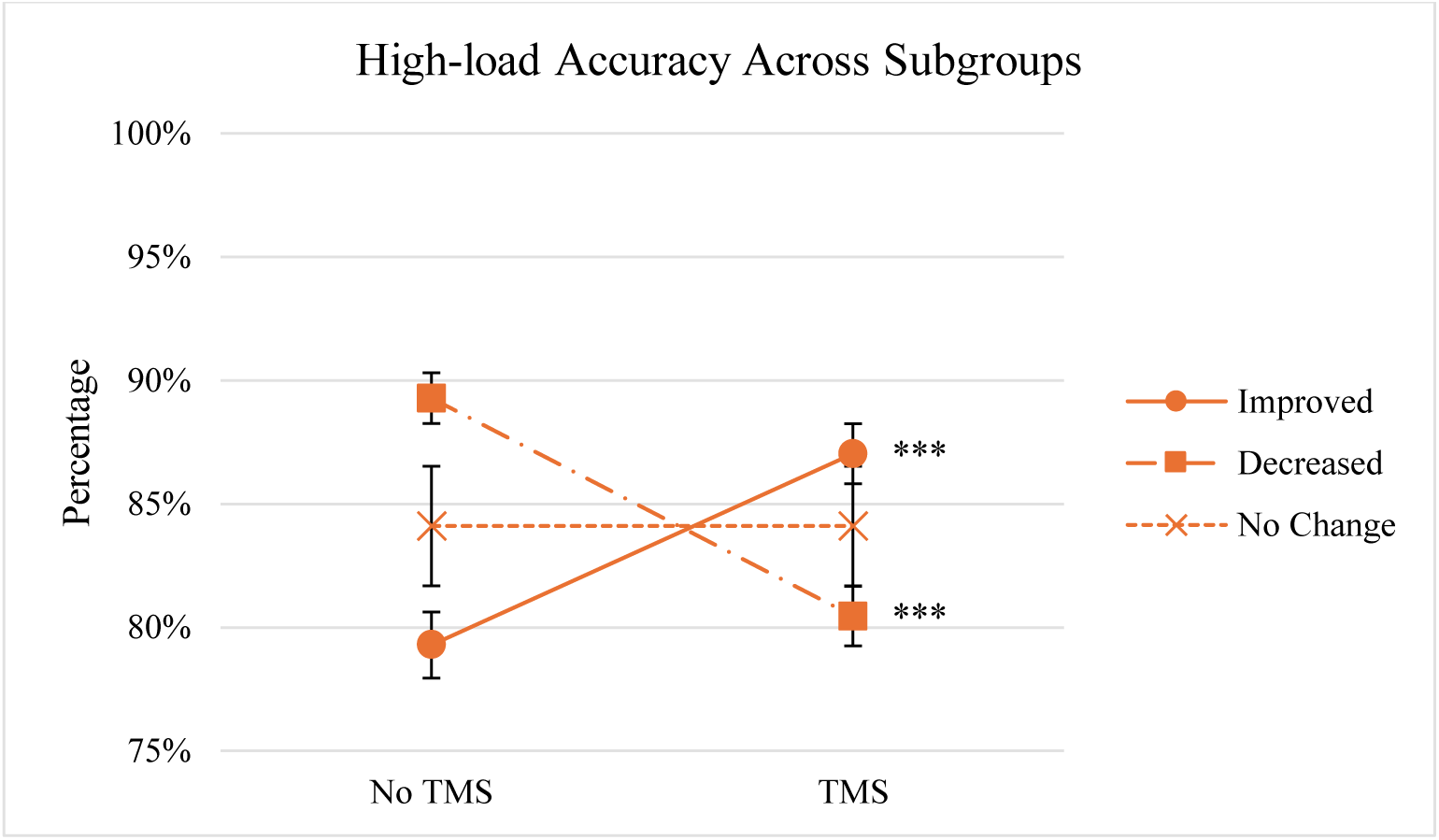
Average accuracy during high-load condition across TMS improved, decreased, and no change subgroups. There is a significant increase in accuracy by TMS in the improved group, and a significant decrease in accuracy by TMS in the decreased group. *(*** p < 0 .001)*. Bars indicate standard error.

**Table 2:**
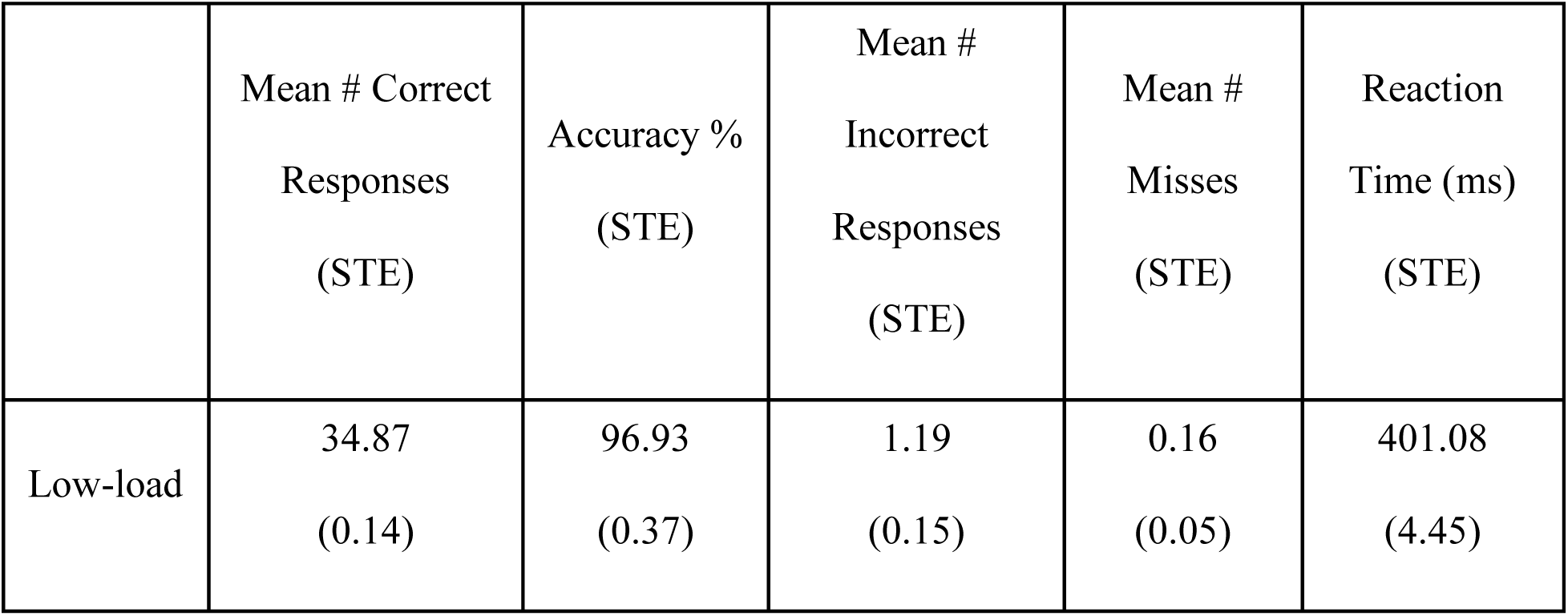
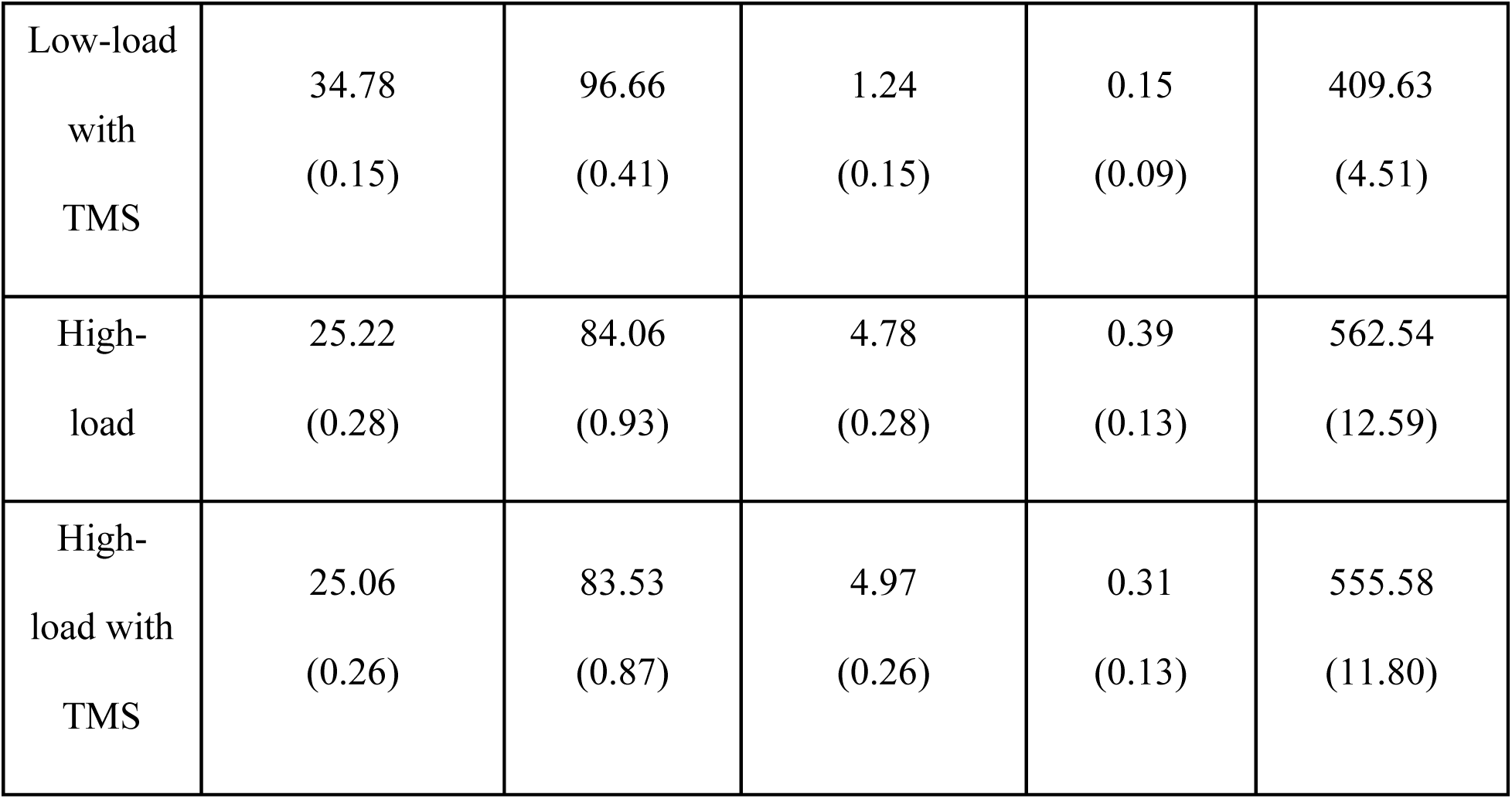
Behavior Results.

|  | Mean # Correct<br>Responses<br>(STE) | Accuracy %<br>(STE) | Mean #<br>Incorrect<br>Responses<br>(STE) | Mean #<br>Misses<br>(STE) | Reaction<br>Time (ms)<br>(STE) |
| --- | --- | --- | --- | --- | --- |
| Low-load | 34.87<br>(0.14) | 96.93<br>(0.37) | 1.19<br>(0.15) | 0.16<br>(0.05) | 401.08<br>(4.45) |
| Low-load<br>with<br>TMS | 34.78<br>(0.15) | 96.66<br>(0.41) | 1.24<br>(0.15) | 0.15<br>(0.09) | 409.63<br>(4.51) |
| High-load | 25.22<br>(0.28) | 84.06<br>(0.93) | 4.78<br>(0.28) | 0.39<br>(0.13) | 562.54<br>(12.59) |
| High-load with<br>TMS | 25.06<br>(0.26) | 83.53<br>(0.87) | 4.97<br>(0.26) | 0.31<br>(0.13) | 555.58<br>(11.80) |

### fNIRS Measured Hemodynamic Responses

Based on the processed fNIRS HbO data, hemodynamic responses of load and TMS effects were estimated by GLM and tested at whole group level. Across all subjects, there are significant increases of hemodynamic activity during high-load with and without TMS compared to low-load, specifically in middle frontal gyrus, inferior frontal gyrus, inferior parietal gyrus, dorsolateral prefrontal cortex, superior medial frontal gyrus, precentral gyrus and middle temporal gyrus (shown in *Figure 4*). No regions were found with significant decreases of hemodynamic activity. The regional peaks of the load effect are listed in *Table 3*.

**Fig. 4.**
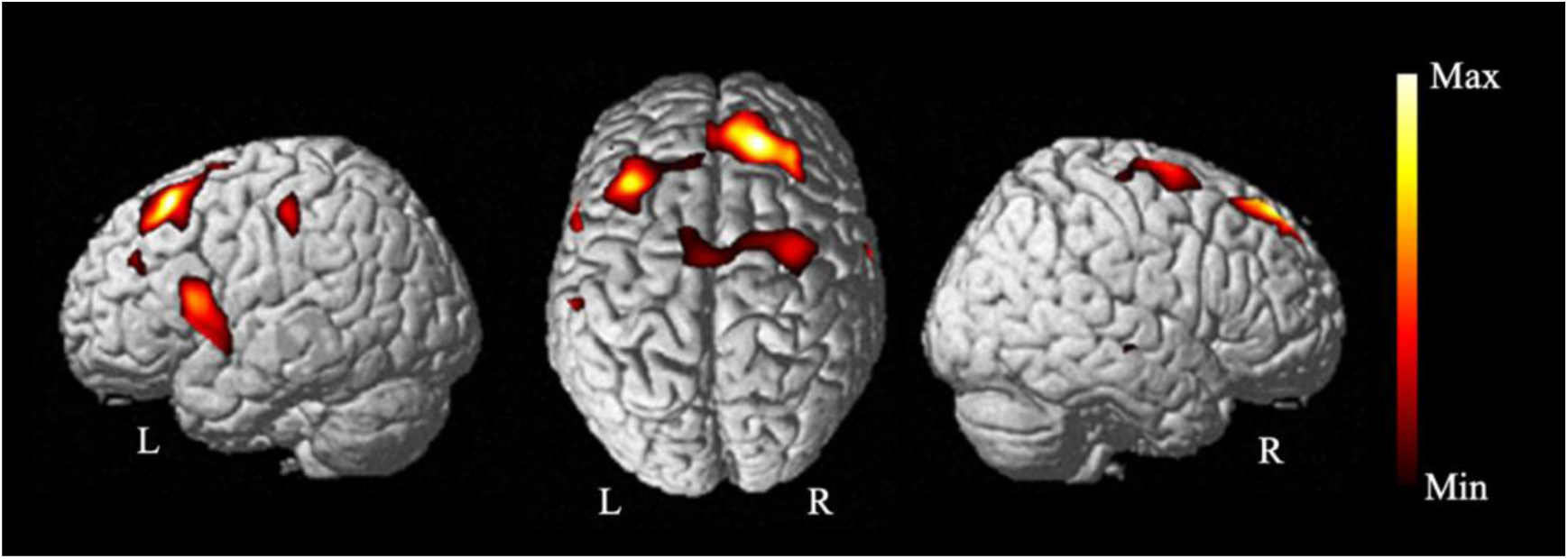
Load effect at the group level. This contrast compared hemodynamic response between high and low working memory load, across TMS and no-TMS conditions. There are significant areas of increases in the left middle frontal gyrus, left inferior frontal gyrus, left inferior parietal gyrus, left dorsolateral prefrontal cortex, right superior medial frontal gyrus, right precentral gyrus and right middle temporal gyrus *(p < 0.05, corrected)*.

**Table 3:**
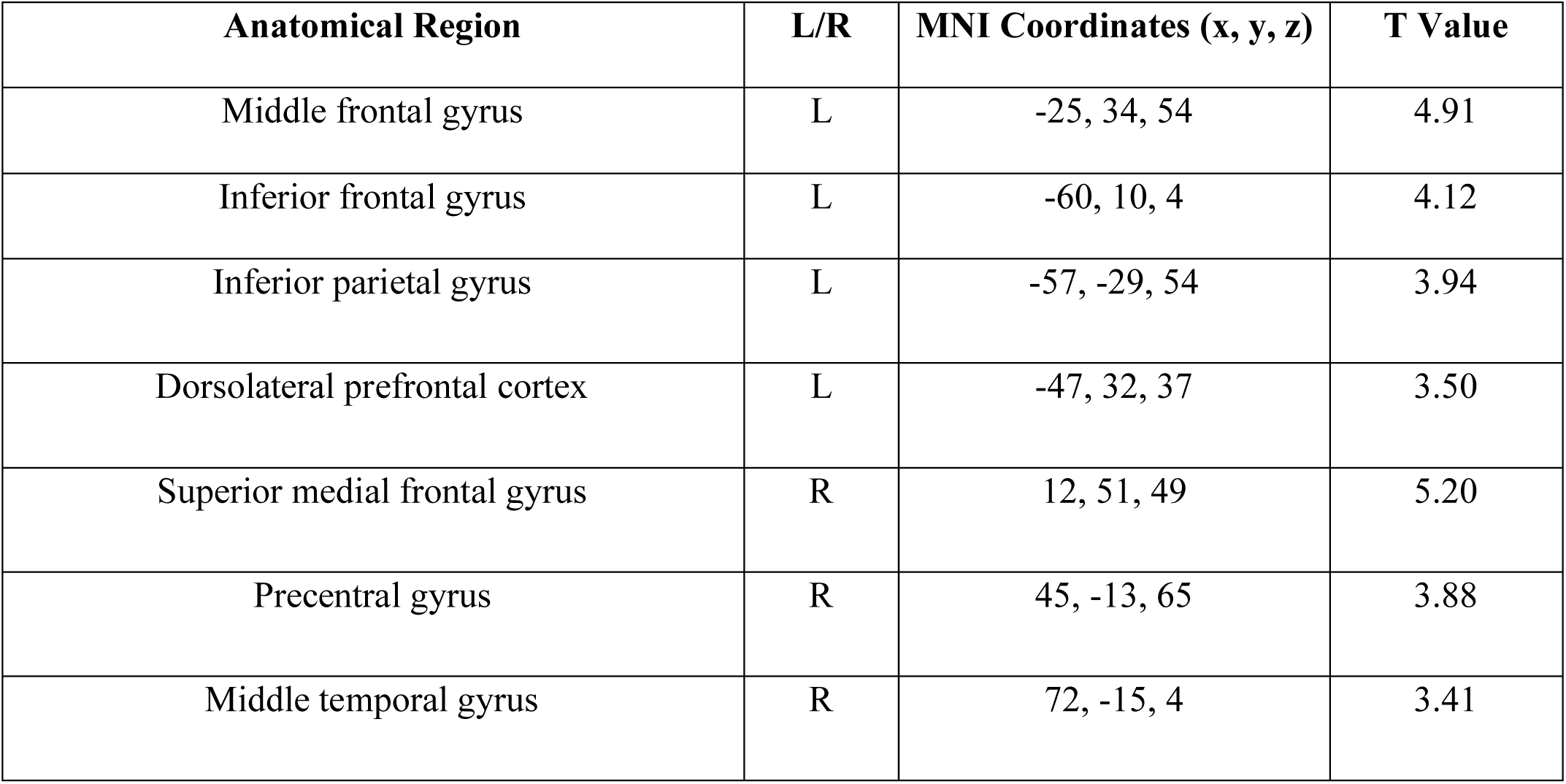
Load Effect Coordinates and T Values.

We then examined the TMS effect at the group level. Like the accuracy results, no significance was found in the whole group; however, there were indeed differences found between improved, decreased, and no change subgroups by the stratified ANOVA analysis. Regions of significance include the left dorsolateral prefrontal cortex, superior medial gyrus, right superior frontal gyrus, left angular gyrus, precuneus, and right superior parietal lobule (shown in *Figure 5*). For the improved group, the hemodynamic response with TMS effect was increased and at the highest level in every region, while the worsened group, however, showed decreased hemodynamic response in every region. The no-change group showed mixed regional responses exhibiting greatest standard error. The TMS effect on hemodynamic response is significant between improved and decreased subgroups in the left dorsolateral prefrontal cortex, precuneus and right superior parietal lobule (*p* < 0.05, corrected), between improved and unchanged subgroups in superior medial gyrus, right superior frontal gyrus, precuneus and right superior parietal lobule (*p* < 0.05, corrected).

**Fig. 5.**
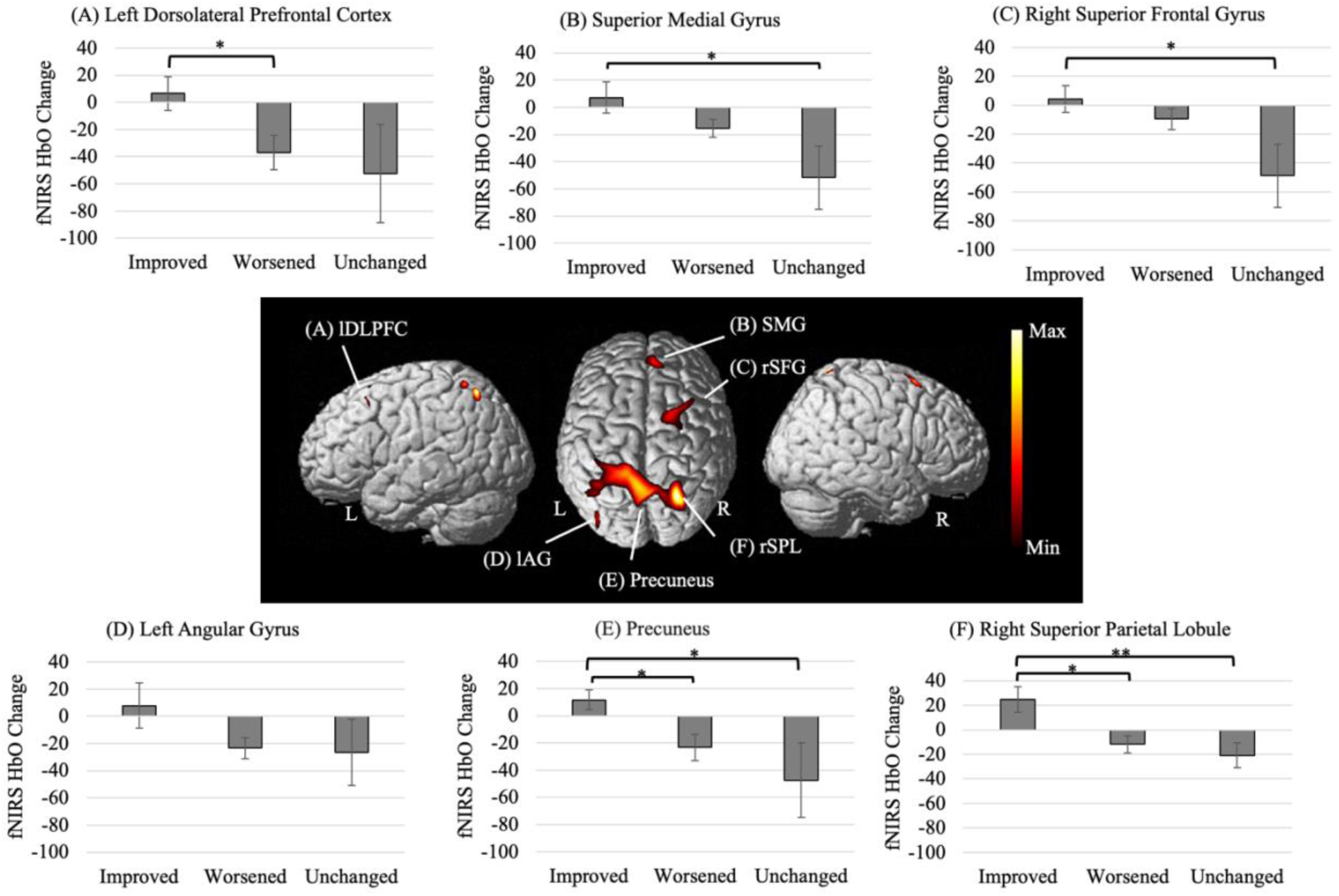
TMS effect is associated with online working memory performance. ANOVA analysis identified areas where hemodynamic responses estimated for the TMS effect are associated with improved, worsened and unchanged performance during working memory task sessions, including (A) left dorsolateral prefrontal cortex (lDLPFC), (B) superior medial gyrus (SMG), (C) right superior frontal gyrus (rSFG), (D) left angular gyrus (lAG), (E) precuneus, and (F) right superior parietal lobule (rSPL) (*p < 0.05*, corrected). Post-hoc comparison found significant differences in activations between improved and worsened subgroups in the lDPLFC, precuneus and rSPL, significance between improved and unchanged subgroups in the SMG, rSFG, precuneus and rSPL. *\** indicates *p < 0.05,* corrected and ** indicates *p* < 0.01, corrected.

**Table 4:** Areas with hemodynamic response associated with TMS improved, worsened and unchanged performance during online task session.

| Anatomical Region | L/R | MNI Coordinates (x, y, z) | F Value |
| --- | --- | --- | --- |
| Precuneus | L | -15, -44, 79 | 4.92 |
| Angular gyrus | L | -38, -72, 55 | 3.86 |
| Dorsolateral prefrontal cortex | L | -42, 20, 56 | 3.43 |
| Superior parietal lobule | R | 29, -58, 71 | 6.27 |
| Superior frontal gyrus | R | 29, 6, 68 | 4.24 |
| Superior medial gyrus | R | 12, 51, 49 | 4.24 |

Furthermore, we tested the TMS effect on the fNIRS measures during single pulses alone without any task. We hypothesized that the variability in the behavior responses TMS is related to the hemodynamic responses as measured by the whole-head fNIRS, regardless that the imaging measures were obtained from separate session without any task. This association, if any, would indicate that the brain responses to single pulse alone could be intrinsically related to or in a certain way predictive of the behavioral responses during an online task manipulation. To explore the association, our analysis performed a stratified ANOVA analysis (improved, decreased and no change subgroups) on the single pulse data that was recorded from the sessions without task. Interestingly, significance related to behavioral responses was found in regions including left dorsolateral prefrontal cortex, precuneus, left/right inferior parietal lobule, and left/right visual cortex (shown in *Figure 6*). The regional peaks are listed in *Table 5*. Especially, hemodynamic response of single TMS pulses in the improved subgroup showed *lower* activities than those in the worsened subgroup, which was observed at both regions of left dorsolateral prefrontal cortex and precuneus. Noteworthy, these responses in the single pulse alone sessions are in an *opposite* direction to those observed during online task where hemodynamic responses of TMS effect in the improved subgroups tend to be higher than in the worsened subgroup. In addition, our analysis tested whether there were significant activations related to single pulses regardless of the behavioral stratification. No activations were found at the level of corrected *p* < 0.05, although decreases of HbO were observed in the left dorsolateral prefrontal cortex, left inferior parietal lobule, and left/right temporal gyrus at uncorrected *p* < 0.05.

**Fig. 6.**
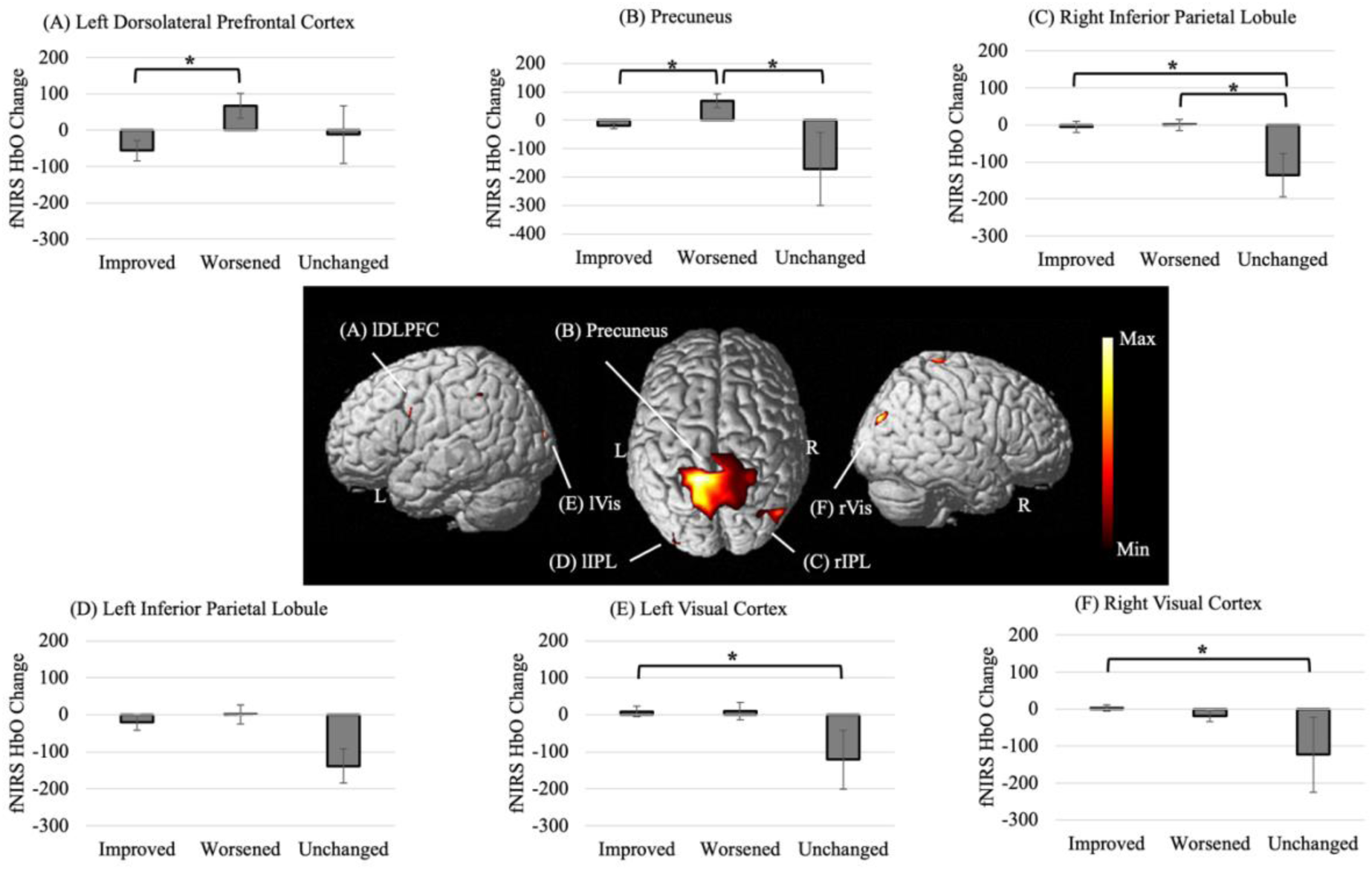
Brain responses to single pulses alone are associated with online working memory performance. ANOVA analysis identified areas where hemodynamic activities during the single pulse alone sessions are associated with improved, worsened and unchanged performance during separate online working memory sessions, including (A) left dorsolateral prefrontal cortex (lDLPFC), (B) precuneus, (C) right inferior parietal lobule (rIPL), (D) left inferior parietal lobule (lIPL), (E) left visual cortex (lVis), and (F) right visual cortex (rVis) (*p < 0.05*, corrected). Post-hoc comparison found significant differences in activations between improved and worsened subgroups in the lDPLFC and precuneus, significance between worsened and unchanged subgroups in the precuneus and rIPL, significance between improved and unchanged subgroups in the rIPL, lVis and rVis. *\** indicates *p < 0.05,* corrected.

**Table 5:** Areas with hemodynamic activity during single pulse alone sessions that are associated with TMS improved, worsened and unchanged performance during online working memory sessions.

| Anatomical Region | L/R | MNI Coordinates (x, y, z) | F Value |
| --- | --- | --- | --- |
| Precuneus | L | -15, -44, 79 | 10.01 |
| Visual cortex | L | -36, -98, 3 | 4.36 |
| Dorsolateral prefrontal cortex | L | -57, 16, 30 | 3.84 |
| Inferior parietal lobule | L | -64, -44, 42 | 3.32 |
| Inferior parietal lobule | R | 53, -70, 37 | 6.81 |
| Visual cortex | R | 46, -82, 28 | 4.84 |

## Discussion

To our knowledge, the present investigation is the *first* online TMS-fNIRS task paradigm to examine how single pulse TMS applied to dorsolateral prefrontal cortex modulates brain activity and working memory performance at various cognitive load. Our results revealed significant findings regarding the effect of cognitive load. Moreover, although behavioral responses by TMS varied, the concurrent TMS-fNIRS measurements during online task revealed significant association between hemodynamic and behavior responses to TMS in local and distant areas. Additionally, hemodynamic response to single pulses in a TMS alone session was also linked to the behavioral responses to TMS regardless of the separate task and no-task sessions, and revealed a coherent topographic pattern.

### Effect of Cognitive Load

Our investigation found that cognitive load modulates the behavioral performance and working memory-related brain activity. Behavioral results revealed an expected load effect, with performance declining as working memory demands increased from low-load and high-load conditions: there is a significant decrease in accuracy between low-load and high-load with and without TMS (results depicted in *Table 2* and *Figure 2*). These behavioral results are consistent with other studies that explored overloading working memory and cognitive effects using fNIRS or fMRI, where accuracy significantly decreased with higher cognitive load (Fishburn et al., 2014; Yun et al., 2010).

Meanwhile, our results revealed a load-dependent pattern in activation across the group of healthy subjects. Specifically, significant areas of activation during high load compared to low load include the superior frontal gyrus, precentral gyrus, middle frontal gyrus, inferior frontal gyrus, inferior parietal gyrus and dorsolateral prefrontal cortex (*Figure 4* and *Table 3*), which are in line with meta-analytic findings of working memory using fMRI without TMS (Owen et al., 2005; Rottschy et al., 2012). These regions, especially DLPFC, inferior parietal gyrus and precentral gyrus, are recognized as belonging to the fronto-parietal network, i.e., FPN (Chen et al., 2013; Fox et al., 2005). Our findings of the working memory associated areas also align with fNIRS literature that reported greater activity in frontal regions during high cognitive load that were investigated without online TMS (Fishburn et al., 2014). In particular, our fNIRS study utilized a whole brain montage and has shown activation pattern involving both frontal and parietal regions of FPN than prior studies that utilized only partial coverage (Fishburn et al., 2014). Moreover, recent studies using fMRI and online TMS reported similar activations in FPN during high cognitive load than low load (Ripp et al., 2021; Webler et al., 2022; Yun et al., 2010). Consistent findings between fNIRS and fMRI support the validity of fNIRS, particularly in its ability to detect key FPN regions. FPN has been indicated to be a common neural substrate involved in working memory as well as repetitive TMS intervention: increasing FPN activation is linked to furthering deactivation of the default mode network, i.e., DMN (Hamilton et al., 2011; Wang et al., 2019; Webler et al., 2022), as well as a weakened connectivity between FPN and DMN (Chen et al., 2013; Liston et al., 2014). Since fNIRS is less expensive and more accessible for use in clinical settings, it could be used to measure activity in working memory-related FPN regions as a proxy for TMS-induced network modulation.

### Effect of Single Pulse TMS

At the group level, our results showed no significant TMS effect, but the behavioral effect of TMS was variable. To account for this variability, we split the data into subgroups based on performance: whether TMS increased, decreased, or had no change than without TMS. In doing so, we examined hemodynamic response across behaviorally defined subgroups to search for the neural substrate. Interestingly, our results revealed that the TMS effect on imaging measurements of brain activity is indeed dependent on the variable behavioral performance: a *brain-behavior association* (*Figure 5* and *Table 4*). Regions of significance between subgroups include the dorsolateral prefrontal cortex, superior medial gyrus, right superior frontal gyrus, left angular gyrus, precuneus, and right superior parietal lobe, which are consistent with those areas reported in prior online TMS-fMRI studies. The middle line regions, especially superior medial gyrus, superior frontal gyrus and precuneus, have been reported to be activated in interleaved TMS-fMRI data by single pulses (Vink et al., 2018), and triplets of single pulses (Tik et al., 2023a; Tik et al., 2023b). Additionally, the superior parietal lobe has also been reported (Vink et al., 2018). These areas activated by TMS delivered towards left DLPFC have been termed as TMS-responsive network (Tik et al., 2023b). However, variability has been noted in prior reports: activations to TMS pulses are not uniformly observed in all subjects of the same study (Tik et al., 2023a; Vink et al., 2018), and even more variable findings across studies (Bergmann et al., 2021). Importantly, our study has further shown that such variability of TMS activations is associated with behavior variability.

Comparing increased, decreased and unchanged behavioral outcomes against the TMS-concurrent imaging measures revealed significant findings. Specifically, the improved group had elevated hemodynamic response while the worsened subgroup exhibited decreased hemodynamic response in every region. The link between higher accuracy in working memory and stronger underlying neural activation, as measured with fMRI or fNIRS, has been consistently reported. Increased accuracy typically corresponds to heightened hemodynamic activity, indicating stronger or more extensive recruitment of brain regions supporting working memory (Zhu et al., 2024). Conversely, disruptions in connectivity and reductions in network efficiency are often observed following TMS, reflecting less effective neural communication in task-relevant areas (Beynel et al., 2020). These network-level inefficiencies can translate into poorer task performance or reduced accuracy. Although our study applied online TMS during the working memory task, the directionality in the observed brain–behavior association aligns with the broader pattern described in prior research without TMS. Thus, greater hemodynamic activity signals enhanced cognitive processing leading to improved performance, while lower hemodynamic activity suggests diminished engagement and poorer accuracy. These results provide valuable insight into how the brain responds to TMS during high cognitive load and align with a growing literature suggesting that TMS treatment may be most effective when administered during working memory tasks, as subjects with higher accuracy and network activation tend to show greater treatment benefit (Sack et al., 2024).

Moreover, in addition to examining hemodynamic response during the online task session, we investigated the fNIRS recordings from the single pulse alone sessions and examined whether a pattern of TMS induced activity aside from the task is intrinsically associated to the behavioral responses. Our analysis showed that TMS-related brain activities during single pulse session without any task are linked to the behavioral responses during online working memory task session. A few regions of brain-behavior-association are commonly identified, including left DLPFC, precuneus and the left/right parietal lobules; these regions are recognized as belonging to the default mode network. The finding suggests that activations to single pulse alone in these regions associated with DMN might be mechanistically linked to behavior responses in separate online TMS manipulations. However, it is important to note that TMS increased hemodynamic response during online working memory, whereas TMS alone decreased hemodynamic response during single pulse alone in these regions. Many prior studies have reported TMS administered during online task do not represent the same pattern of activations during TMS lone (Sack et al., 2024). Our observation of opposite directions of changes suggests state-dependent mechanisms of TMS modulation, especially in the areas that are commonly linked to behavioral responses.

### Explanations of Variability in TMS effect

A few factors may explain the variability seen in our data. The effect of TMS on task behavior and the brain network is known to be variable, depending on when it is applied, brain state, and variability in anatomy or excitability (Beynel et al., 2020; Luber and Lisanby, 2014). The timing of TMS could matter, especially relative to the endogenous cognitive and perceptual state of the subjects. In an online TMS-fMRI working memory paradigm, Grosshagauer et al. manipulated the TMS pulses to be before and after the memory probe and showed that TMS differentially modulate brain networks depending on the timing of cognitive states (Grosshagauer et al., 2024). In addition, the power or phase of underlying oscillatory activities could affect the TMS response. A recent study by Pantazatos et al. revealed the influence of EEG alpha phase during TMS pulses on modulations of subgenual anterior cingulate cortex (sgACC) activity and connectivity (Pantazatos et al., 2023). The timing and the endogenous oscillatory state could drive the variability in our data. However, these studies did not show clear evidence of linking the variable behavior performance to the brain states under TMS. Nonetheless, we have found an association between the behavioral performance and the brain involvement in a topographic manner that is coherent from TMS with online task to TMS without task conditions.

Moreover, variability is reported in DLPFC excitability and anatomy. Although stimulation intensity is often set using resting motor threshold (and was used in this protocol), this method does not appear to generalize well to non-motor regions like the DLPFC. Interleaved TMS-fMRI demonstrates that individuals show significantly different dose-response profiles to DLPFC stimulation, where some exhibit peak activation at sub-threshold levels while other require supra-threshold intensities, suggesting that RMT does not reliably reflect DLPFC excitability (Tik et al., 2023a). Additional resting-state fMRI studies have shown higher inter-individual variability in spatial localization of functionally defined DLPFC targets (Ning et al., 2019). Specific measures find that the average inter-day variability of DLPFC, based on the functional connectivity with subgenual cingulate, exceeds 25 mm which often surpasses the spatial precision of standard TMS coils (Ning et al., 2019). This is especially important to note since we targeted based on BEAM-F3 method and did not consider the individuals’ functional connectivity. Even though anticorrelation with sgACC has been strongly suggested to be optimized for repetitive TMS treatment (Cash et al., 2021), there is no consensus in personalizing the target selection for enhancing working memory performance. Grosshagauer et al. (2024) has selected their target based on the sgACC anticorrelation, but they could not detect significant behavioral effect either. Nonetheless, our results have linked a distinct pattern of hemodynamic responses to the variable behavioral performance during online TMS task sessions. Our exploratory finding suggests a possible strategy to personalize the TMS target - maximizing the fNIRS measured hemodynamic response in a few specific brain areas, which may lead to more consistent behavioral effect.

Additionally, TMS response could be dependent on the brain state of subjects. Webler et al. (2022) reported TMS enhancement of high-load performance at the group level, while their average high-load accuracy was at 63.70% without TMS. Noteworthy, our study had a high-load accuracy of 84.26% without TMS (results depicted in *Table 2* and *Figure 2*) and the Grosshagauer et al. study (2024) had a baseline high-load accuracy of 97.40%, while neither had a group-level enhancement of high-load performance with TMS. It is possible that when baseline performance started so high, TMS would not be able to increase accuracy enough to be significant. The interaction between TMS effects, cognitive ability and task difficulty has been shown previously (Heinen et al., 2011; Sack et al., 2007; Silvanto et al., 2017). Correspondingly, our population had an average education of 16.85 years (results depicted in *Table 1*). Literature supports this theory since people with more schooling may perform better on high cognitive load tasks because education enhances cognitive skills like attention, processing speed, and memory strategies (de Souza-Talarico et al., 2007). However, when probing the variability in a clinical population, possibly with impaired performance than normal, whether the TMS could improve the performance is yet to be seen.

### Clinical Implications

Our neuroimaging findings showthat fNIRS may be used to assess the fronto-parietal network, especially based upon the FPN associated regions identified via the load-dependent effect (Beynel et al., 2020; Herbet et al., 2016). This fNIRS approach has promising implications for assessing working memory in both healthy and clinical populations and delineating the neural circuits that may be key to depression interventions (Luber and Lisanby, 2014; Zhu et al., 2024).

Additionally, our results showed that the single pulse related fNIRS activity has been linked to the behavioral effect of TMS. The results support the interpretation that TMS effects on working memory are highly variable. Furthermore, the variability in TMS-induced behavioral and hemodynamic responses seen in healthy subjects likely reflects even greater variability in patients with neurophysiological disorders like depression, whose neural architecture may be dysfunctional.

Recognizing the variability in TMS response is critical for tailoring neuromodulation interventions, as a one-size-fits-all approach may fail to capture the variability in network plasticity and target engagement (Soleimani et al., 2025). Our findings are exploratory, but they are also unique and significant in highlighting meaningful variability in brain activity that is associated with behavioral outcome. Such a discovery suggests a possible strategy to personalize the TMS target - maximizing the fNIRS response in a few specific brain areas in order to enhance the behavioral performance during online TMS. In a future experimental design, for example, concurrent fNIRS-TMS may recognize regions with heightened TMS affects in real time to deliver stimulation at an optimal site or a preferred state during a working memory task.

### Limitations

This study has several limitations that should be considered when interpreting the results. We did not include auditory or sensory sham TMS conditions, which limits our ability to fully isolate TMS-specific effects from non-specific stimulation or task-related influences. Although we partially addressed this by contrasting differences between TMS and without TMS conditions as well as examining differences across behavioral response subgroups as an internal comparison, this does not fully substitute for an external control condition.

Also, our targeting approach was based on the Beam-F3 method. Given known inter-individual variability in DLPFC anatomy and functional organization, this may have introduced variability in stimulation target. Additionally, the stimulation intensity in our study was calibrated using resting motor threshold, which may not accurately reflect excitability in non-motor regions such as the DLPFC. This could have led to variability in the effective dose delivered at the stimulation site and contributed to variable behavioral and hemodynamic responses. We did not include electric field (E-field) modeling, which limits our ability to identify the factors contributing to the variability observed in our data, including coil placement, head anatomy, tissue conductivity etc. However, future study is warranted to model the variability of target and dose, and further determine whether controlling the precision of stimulation could control the variability.

Finally, our sample consisted of relatively high-functioning and highly educated subjects, which may have resulted in ceiling effects in task performance and reduced sensitivity to detect behavioral modulation by TMS.

## Conclusions

Our findings provide novel insights into how single-pulse TMS combined with fNIRS can reveal the mechanism of working memory function in healthy subjects using a very unique online TMS paradigm with concurrent whole-head fNIRS imaging. A consistent load-dependent increase in fNIRS measured hemodynamic response was observed in regions associated with the FPN, confirming a network-level neural engagement during working memory. Additionally, although the TMS effect on the behavioral performance was found to be variable, our stratified analysis of improved, decreased, and no-change responses revealed a distinct topographic pattern of hemodynamic response that is associated with the behavioral responses, including the dorsolateral prefrontal cortex, superior medial and frontal gyrus, precuneus, angular gyrus and superior parietal lobule that are recognized as key regions of the default mode network. Moreover, the hemodynamic activity to single pulses alone without any task also exhibited a coherent pattern that is associated with the behavioral responses. Our findings suggest that variable behavioral outcomes during online TMS task are linked to distinct hemodynamic responses in a topographic pattern of local and distant regional areas. Although the variability in behavioral responses to TMS has been acknowledged in many studies, to our knowledge, no prior research has linked the TMS-concurrent imaging measures during online TMS task as well as the during single pulses alone to the behavioral response, making these results both unique and a significant contribution to understanding variability in brain stimulation outcomes. Importantly, our results underscore the importance of further characterizing the variability and suggest a potential strategy to achieve consistent cognitive behavioral outcomes. In the future, our findings from concurrent TMS-fNIRS imaging may have relevance to clinical populations, as they could potentially improve the treatment efficacy via personalizing stimulation parameters based on real-time imaging.

## Declarations

## Author Contributions

SE: Investigation, Formal analysis, Methodology, Writing - original draft. QS: Investigation, Formal analysis, Methodology, Writing - original draft. JF: Investigation, Formal analysis, Methodology, Writing - original draft. TMS: Investigation, Formal analysis, Writing - original draft. LD: Writing - original draft. AKC: Conceptualization, Funding acquisition, Writing - original draft, Supervision. IFD: Conceptualization, Funding acquisition, Writing - original draft, Supervision. MSG: Conceptualization, Methodology, Writing - original draft. HY: Conceptualization, Methodology, Formal analysis, Funding acquisition, Supervision, Writing - original draft.

## Data Availability Statement

Data used in this study are not publicly available due to data sharing restrictions from the IRB but are available from the corresponding author through a data use agreement upon reasonable request.

## Acknowledgements

We thank the staff at the Department of Neurosurgery and the Neurosurgery Clinic at University of Oklahoma Health Campus for their assistance in data collection.

